# Impact of social media on knowledge dissemination between physicians during COVID-19 virus outbreak: A cross sectional survey

**DOI:** 10.1101/2020.05.31.20118232

**Authors:** Aliae AR Mohamed-Hussein, Nahed A Makhlouf, Heba A Yassa, Hoda A Makhlouf

## Abstract

**Background:** Social media became an alternative platform for communicating during medical crisis as COVID-19 pandemic.

**Aim of the study:** 1- to describe the use of social media by Physicians during Covid-19 outbreak 2- to determine how physicians obtain their medical information about the emerging disease 3- to determine physicians practice and how do they use the information received.

**Methods:** This is a cross-sectional web-based anonymous survey. Data were collected from Health Care Professional (HCPs) via fulfilling online designed questionnaire. Descriptive statistics with frequencies and percentages are presented. **Results:** The response rate was 66.2% (232/350). Smart phones was the most commonly used (94.8%) followed by laptops (13.4%). Facebook was used by 65.8% and WhatsApp by 52.8%. The data shared were medical newsletters (68%) and educational movies (52.2%). Source of information were mainly professional local pages (60.8%) then WHO pages (53.7%). Physicians shared trusted information (66.7%) and they confirmed the data were correct before publishing in 55.5%. They shared mainly WHO announcements and alerts (44%), professional lectures (32.1%) and 13.3% shared comics. Overall, 71% perceived lots of data about the cause of disease, clinical picture, daily spread, fatality rate and alert of the countries.

**Conclusion:** Physicians are active users of social media. Facebook and WhatsApp are useful platforms to spread right data about diseases during pandemics. Most physicians are positive towards data published; they watch, read and disseminate trusted informations.

## Introduction

The past few years have seen a boom of social media such as Facebook, WhatsApp, LinkedIn, Twitter and other newer methods of dissemination (1). Social media has the ability to reach a large audience in a short time, which leads to fast communication (2). It is assumed that physicians are the first line of defense (White Army) against the COVID-19 spread, most of people tried to take their data from their doctors to avoid false data that may cost lives among people in the community.

In December 2019, cases of pneumonia of unknown etiology were detected in Wuhan (3), and a new coronavirus, called SARS-CoV-2 was extracted from lower respiratory tract samples of several patients. The infection spreads to different countries of the world. As of March 10, 2020, more than 100,000 cases were diagnosed worldwide (4). As of March 11, 2020, the WHO has declared the infection a Pandemic, which indicates significant worldwide spread of the disease (5). On April 3, 2020, the WHO has reported that, there are more than 1 million confirmed cases of COVID-19 including more than 50,000 deaths (6).

Social media is useful for spreading medical information, but the quality of the provided data varies and may be inaccurate and misleading (7, 8). Whether specific type of social media may be more preferred to another is not known.

### This study aimed to

1- describe the use of social media by Physicians during COVID-19 outbreak. 2- Determine how Physicians obtain their medical Information about emerging diseases. 3- And assess Physicians practice and how do they use the information received.

## Design and Methods

### Design

This was a cross-sectional web-based anonymous survey.

### Population

Data were collected from participants via fulfilling online designed questionnaire taking in consideration data confidentiality. Participants were invited to complete the survey through email and whatsapp campaign from 1^st^ march to 1^st^ April 2020.

### Sampling

Convenience sampling was used to contact as many Health Care Professional (HCPs) as possible over 1 month. They completed the online questionnaire.

### Measures

We developed and validated the questionnaire used in this study. After establishing face and content validities, internal validity was established in a pilot study of 30 participants. Cronbach was found to be 0.7. We used Google forms for designing the online questionnaire. The questionnaire consists of 2 parts: demographic characteristics and attitude questions. Demographic characteristics questions including age (in years), gender (male or female), job category and country. Attitudes of HCPs in the online Questionnaire include used sources of information, type of messages received or shared by the responding physicians and Physicians practice and how do they use the information received.

### Ethics

The participation of participants was optional, and the response of each individual was confidential.

The clinicalTrial ID: NCT04319315

### Statistical Analyses

Data were analyzed using Google sheets form. Data are expressed as number (%) as appropriate.

## Results

The response rate to the survey was 66.2% (232/350). Table 1 shows the demographic data of the physicians who responded to the study. Male/female ratio was 32.8/67.2% with age ranged between 25–65 years old, most of them from Egypt (67.7%). They are of different level of profession; most of them were medical specialist and consultants (40.7% and 34.1% respectively).

**Table 1.** Demographics data of the included responding physicians (n = 232)

|  | Responders | Number | Percentage % |
| --- | --- | --- | --- |
| **Gender (male/female)** | **232** | -- | **32.8/67.2** |
| **Age ( years/old)** | **227** |  |  |
| 25-35 |  | 72 | 31.7 |
| 36-45 |  | 40 | 17.6 |
| 46-55 |  | 90 | 39.6 |
| 56-65 |  | 25 | 11 |
| **Country- residence** | **229** |  |  |
| Egypt |  | 155 | 67.7 |
| KSA |  | 12 | 5.2 |
| UAE |  | 6 | 2.6 |
| Other |  | 56 | 24.5 |
| **Current job** | **226** |  |  |
| Specialist |  | 92 | 40.7 |
| General practitioner |  | 42 | 18.6 |
| Consultant |  | 77 | 34.1 |
| Other healthcare provider |  | 16 | 7.1 |

KSA= Kingdom of Saudi Arabia, UAE= United Arab of Emarates

Table 2 demonstrates the different sources used by physicians to spread information. Most of them used smart phones by 94.8%, followed by laptops (13.4%). Time spent on the Internet varies from <1 hour to > 3 hours, and the most used platform to spread information was Facebook by 65.8% followed by WhatsApp by 52.8%. The data shared by the Physicians are described in Table 3. These were mainly medical newsletters (68%) and educational movies (52.2%). Type of pages used as a source of information were mainly professional local pages (60.8%), WHO pages (53.7%) and some used family and friends’ pages (48.5%).

**Table 2.**
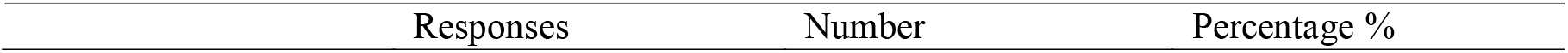
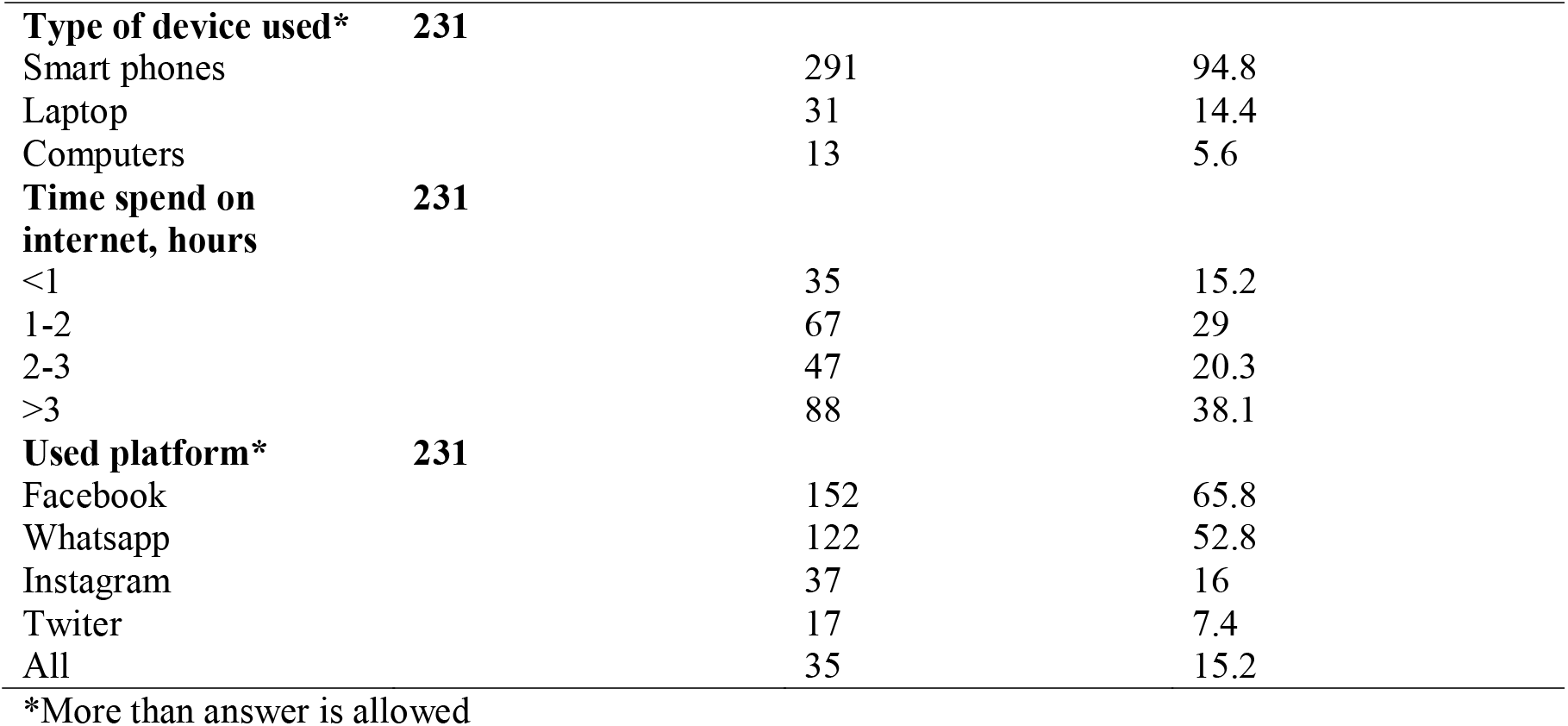
Used sources of information by responding physician included in the survey (n = 232)

**Table 3.** Type of messages received or shared by the responding physicians included in the survey (n = 232)

|  | Responses | Number | Percentage |
| --- | --- | --- | --- |
| **Received messages\*** | **228** |  |  |
| Educational movies |  | 119 | 52.2 |
| Photos |  | 83 | 36.4 |
| Medical newsletters |  | 155 | 68 |
| Unknown source informations |  | 66 | 28.9 |
| Email |  | 43 | 18.9 |
| **Type of pages used as a source of information\*** | **227** |  |  |
| Professional local pages |  | 138 | 60.8 |
| WHO pages |  | 122 | 53.7 |
| Non professional pages |  | 51 | 22 |
| Family and friends pages |  | 110 | 48.5 |

- More than option could be selected

Table 4 illustrates the practice of Physicians towards information they received, they mainly read and share trusted information (66.7%) and they 55.5% confirm the data before publishing, only 8.8% do not confirm the information before spread.

**Table 4.**
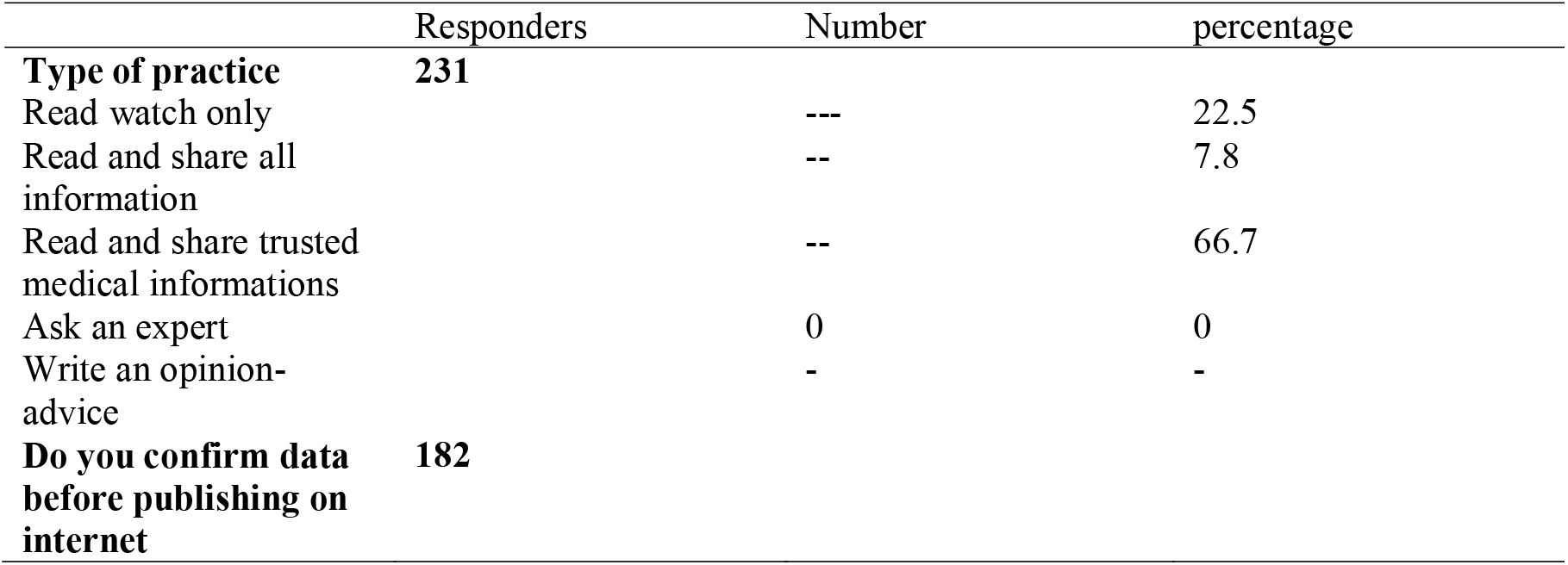
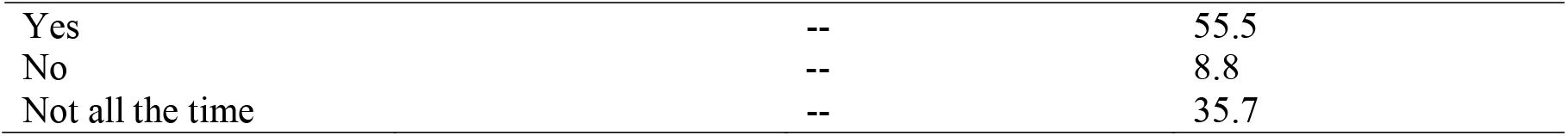
Physicians practice and how do you they use the informations received (n = 232)

|  | Responders | Number | percentage |
| --- | --- | --- | --- |
| **Type of practice** | **231** |  |  |
| Read watch only |  | --- | 22.5 |
| Read and share all information |  | -- | 7.8 |
| Read and share trusted medical informations |  | -- | 66.7 |
| Ask an expert |  | 0 | 0 |
| Write an opinion-advice |  | - | - |
| **Do you confirm data before publishing on internet** | **182** |  |  |

|  |  |  |
| --- | --- | --- |
| Yes | -- | 55.5 |
| No | -- | 8.8 |
| Not all the time | -- | 35.7 |

Figure 1 demonstrates the type of shared knowledge which was mainly WHO announcements and alerts (44%), professional lectures (32.1%), however, 13.3% felt that they have to share comics.

**Figure 1.**
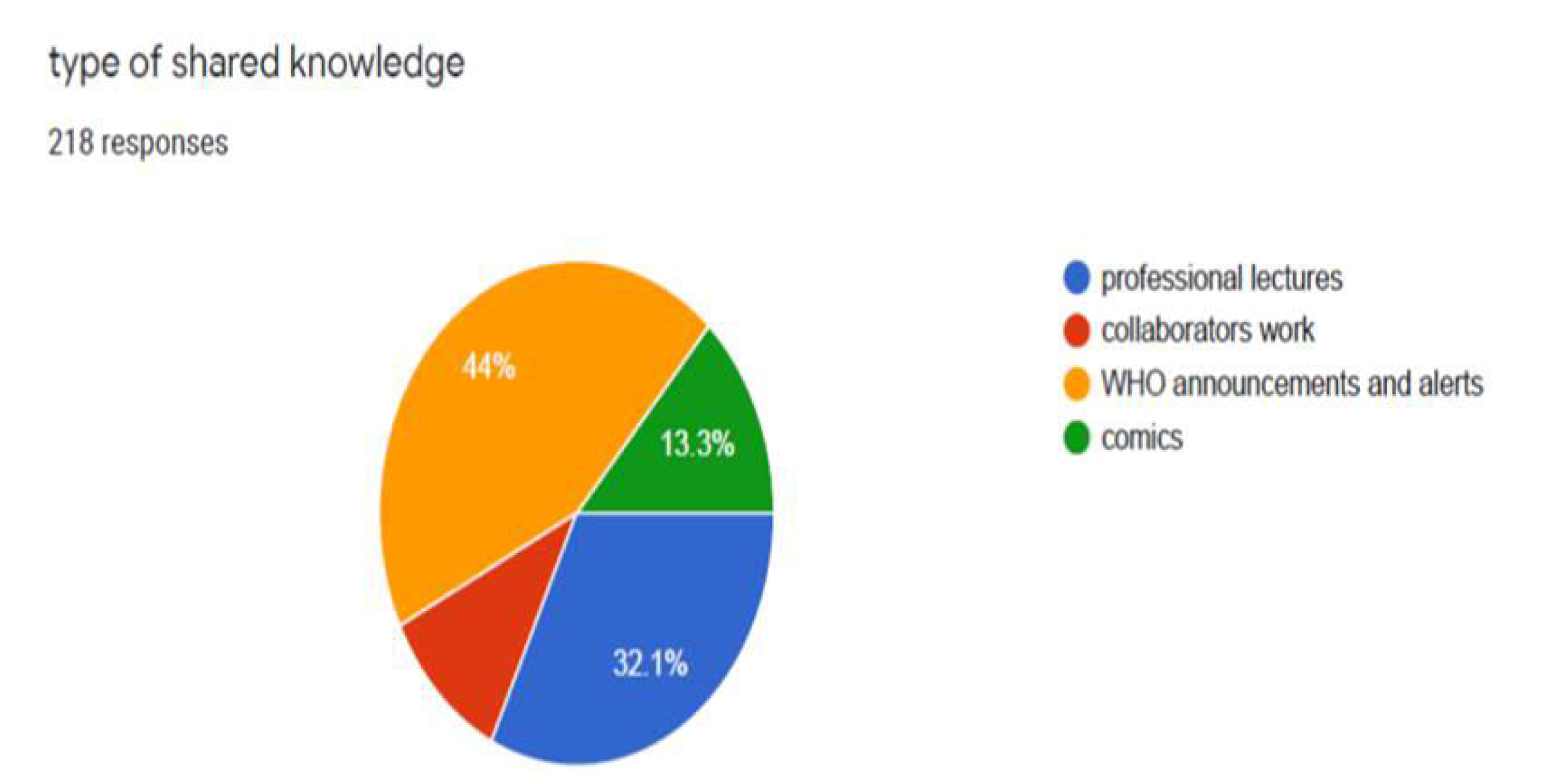
Type of shared knowledge received and shared by Physicians responded to the survey (n=232)

Figure 2 shows that the type of knowledge used by the Physicians shared in the study. Most of them used researches, interdisciplinary collaboration local health services and online training courses (32.3, 29.1, and 13.6% respectively). Lastly, Figure 3 shows the most common perceived knowledge learnt from social media about COVID-19, 71.8% browsed data on all aspects of the disease, including infection control, cause of disease, clinical picture, daily spread, fatality rate and alert of the countries.

**Figure 2.**
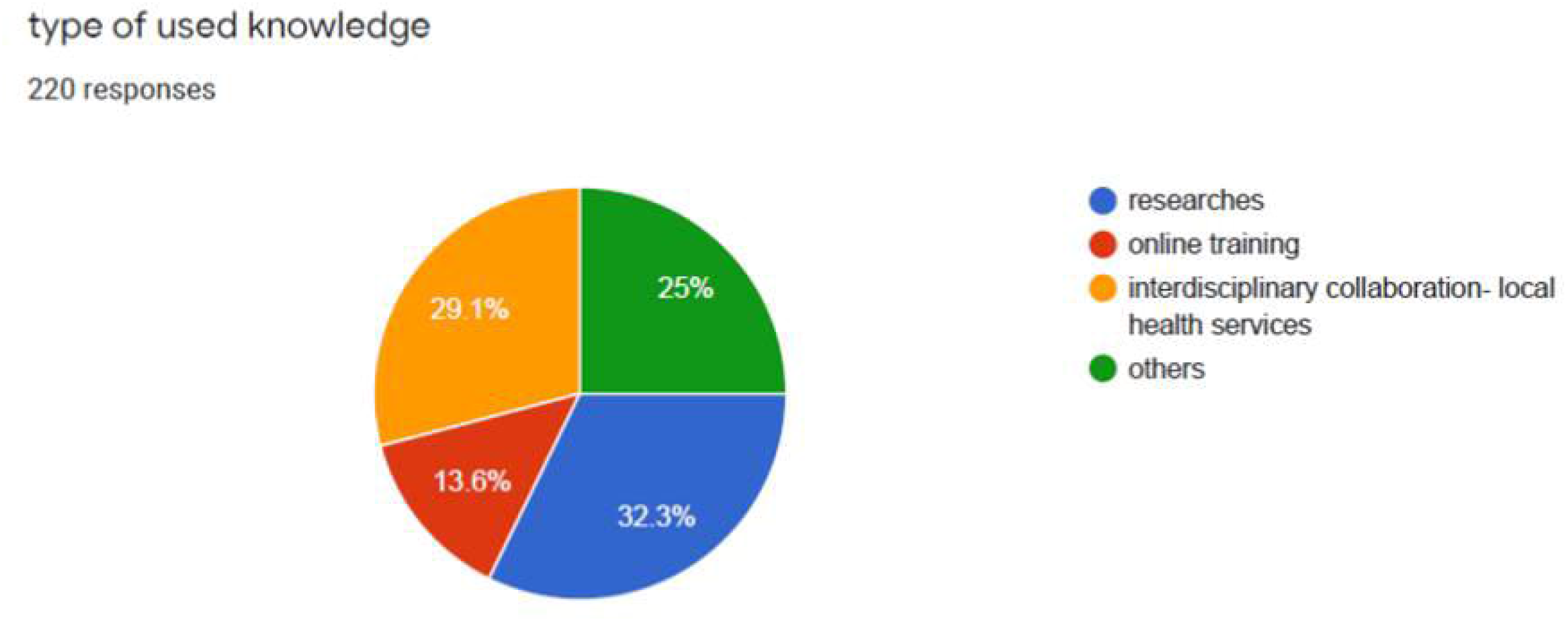
Type of used knowledge by Physicians who responded to the survey (n=232)

**Figure 3.**
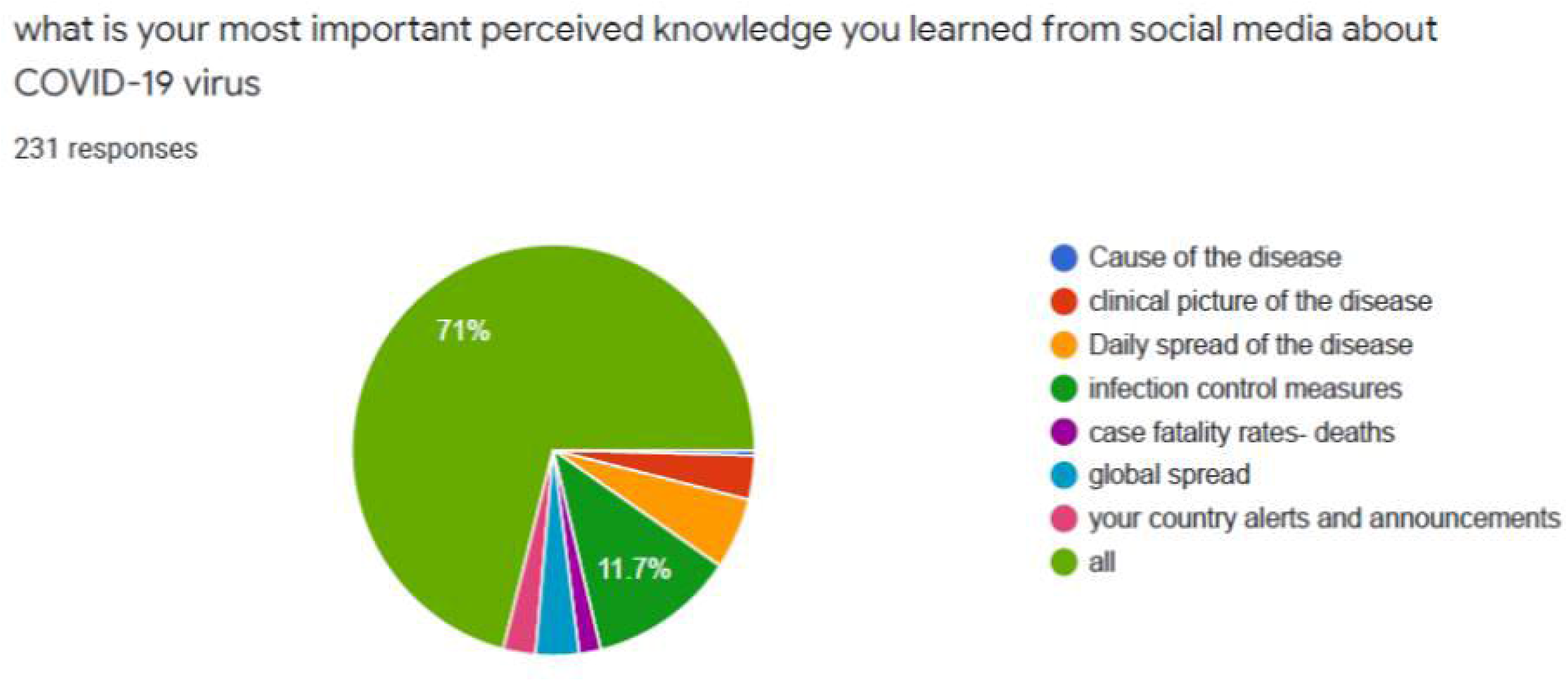
What is the most important knowledge searched and shared by Physicians who responded to the survey (n=232)

## Discussion

The present cross-sectional survey aimed to describe the use of social media by physicians during COVID-19 outbreak. It showed relatively high response rate, dominant use of Smart phones exceeding Laptops and computers, and preference of Facebook and WhatsApp as platforms. Physicians tended to read and share medical newsletters and educational movies (52.2%) and used professional local pages and WHO pages as main source of data. They shared trusted information (WHO announcements and alerts, professional lectures) and usually confirmed the data were correct before publishing in 55.5%. They were concerned about all aspects of the COVID-19 pandemic (cause, spread, death rate, infection control).

The findings of active response to social media between physicians are described in many studied including most of healthcare providers as intensivests, radiologists, emergency medicine doctors, and even participants of continuing medical education courses. Participants (9, 10, 11).

Rumors as social recognition for information, is the fuel that can fire fear among the community. Rumors are usually about great or major issues that have significance among the community (12). Nowadays, social media becomes one of the main sources of spreading information because of its wide dissemination and its use by all levels of community. On the other hand, it is the main source of spreading false information about great issues especially health issues (13). The main issue of this paper is that to know how physicians received the right and wrong data on the emerging disease (COVID-19) and their attitude in spreading information about this great health issue. Most of Physicians used smart phones in their work, as their work in hospitals make the use of smart phones the most easily one to be handled. This explanation accepted by many authors (14).

The majority of data about Coronavirus (COVID 19) shared on the internet comes from fake sites (15). The cause that people spread rumors may be due to feel superior, to attract attention or to feel that they have the power. But all of these reasons not present in most of the doctors that are already having the power and attraction nowadays in the period of COVID 19 spread. In the present study, most of physicians searched for medical newsletters and educational movies to take information that can use to spread it by internet, from WHO sites as well as professional lectures (44.3 and 32%) and 55.2% always confirmed the data before spread. It is known that the Physicians react differently towards rumors and information that spread in their pages. They know well that misinformation and fake news mean that bad advice can circulate rapidly and also can change the reactions of people towards the great risks (16).

## Conclusion

Physicians are active users of social media. Facebook and WhatsApp are useful platforms to spread right data about diseases during pandemics. Most physicians are positive towards data published; they watch, read and disseminate trusted informations.

### Recommendations

Physicians are the first line of defense (White Army) against the COVID-19 spread, most of people tried to take their data from their doctors. Physician should be more active by spreading their information from trusted sites because they know well that false rumors could cost lives among people in the community.

## Data Availability

Data is available on request

## Acknowledgment

The authors want to thank Dr. Suha Baloushah, Reproductive health department, Nursing and midwifery school, Gaza strip, Palestine and Dr. Safa Elrais, University of Tripoli, Faculty of Medicine, Libya, for their active participation in distributing the survey in their institutes.

## Tables

**Table 1.** Demographic data of the included responding Physicians (n = 178)

**Table 2.** Used sources of information by responding Physicians included in the survey (n = 178)

**Table 3.** The type of messages received or shared by the responding Physicians included in the survey (n = 178)

**Table 4.** Physicians practice and how do they use the information received (n = 178)

